# Lecanemab and Anticoagulants: Projected Effects on Health and Quality of Life

**DOI:** 10.1101/2025.04.03.25325187

**Authors:** Sachin J. Shah, Tai Dinger, Deborah Blacker, Steven M. Greenberg, John C. Giardina, Jacquelyn M. Lykken, Samhita Kalidindi, Livia Qoshe, Joseph P. Newhouse, Ankur Pandya, John Hsu, Emily P. Hyle

## Abstract

**Background:** Lecanemab slows cognitive decline among people with early Alzheimer’s disease (early AD) but appears to increase the risk of intracranial hemorrhages (ICHs), including anticoagulant-related ICHs.

**Objective:** To examine the benefits and harms of co-prescribing lecanemab and anticoagulants in people with atrial fibrillation (AF) experiencing early AD.

**Design:** Microsimulation model to compare four treatment strategies. Using inputs from the literature, we modeled increased ICH risk with lecanemab (2.02-fold), apixaban (1.84-fold), and lecanemab/apixaban interaction (2.67-fold). We assigned quality-of-life estimates and increased mortality risk with cognitive decline, stroke, and ICH.

**Data Sources:** Clinical trials, observational cohorts

**Target Population:** People 65-90 years with AF and early AD

**Time Horizon:** 18-month

**Intervention:** Apixaban (*APIX*), apixaban and lecanemab (*APIX/LEC*), lecanemab (*LEC*), neither

**Outcome Measures:** ICH, ischemic stroke, cognitive decline, quality-adjusted life months (QALMs), and survival, age-stratified.

**Results of Base Case:** For 100,000 simulated persons aged 65-74 years, *APIX*, *APIX/LEC*, and *LEC* would result in a similar clinical benefit (13.2 QALMs). Compared to *APIX*, *APIX/LEC* would result in more ICH events (1,990 vs. 400), all-cause deaths (5,820 vs. 5,140), but slower cognitive decline (mean CDR-SB change, 1.11 vs. 1.53). For persons ≥75 years, *APIX* alone would always be preferred.

**Results of Sensitivity Analysis:** Results are sensitive to lecanemab-anticoagulant interaction on ICH, baseline ICH risk, and lecanemab’s effect on cognition.

**Limitations:** Significant parameter uncertainty; treatment burden and costs were not modeled.

**Conclusions:** Model-based results support anticoagulants alone as the preferred strategy for people ≥75 years with early AD and AF. There was greater equipoise across treatment strategies for persons 65-74 years, for whom improved estimates of the ICH risk and lecanemab-anticoagulant interaction are critical to identifying the preferred strategy.

**Primary Funding Source:** National Institute on Aging/National Institutes of Health (K76AG074919, P30AG062421, U01AG076478, and R01AG069575).

## INTRODUCTION

Lecanemab is the first anti-amyloid monoclonal antibody to receive full FDA approval for slowing cognitive decline in people with mild cognitive impairment and mild dementia due to Alzheimer’s Disease (early AD). In Clarity AD, the phase III trial that assessed its efficacy, lecanemab slowed cognitive decline by an average of 0.45 points on the Clinical Dementia Rating-Sum of Boxes (CDR-SB) score, or 27%, over 18 months (1). Regulatory approval from the Food and Drug Administration (FDA) and coverage from the Centers for Medicare and Medicaid Services (CMS) have excited both patients and physicians as a potentially disease-modifying therapy for AD. However, enthusiasm for lecanemab has been tempered because the degree of cognitive improvement may not be clinically meaningful, and it has potentially serious adverse events (1–7).

Intracerebral hemorrhage (ICH), a severe adverse event associated with lecanemab use, is particularly worrisome because of the associated disability and death. In secondary analyses of Clarity AD, concurrent anticoagulant use was among the most potent predictors of ICH (8). In an analysis of randomized data, lecanemab appeared to increase the incidence of ICH among all participants, particularly among those concurrently using anticoagulants, suggesting a possible interaction between lecanemab and anticoagulants (9). Based on these results, expert guidance from the Alzheimer’s Disease and Related Disorders Therapeutics Work Group recommended against the concurrent use of lecanemab and anticoagulants (9). These findings, however, resulted in neither an FDA label warning nor a CMS coverage restriction (10,11).

The lecanemab-anticoagulant drug combination is salient because a concurrent indication for both lecanemab and anticoagulants is common in older adults. For example, atrial fibrillation (AF), the most common indication for indefinite anticoagulation in older adults, co-occurs in 12% of the estimated 6.5 million people with AD in the US (12–14). Because of the greater risk of ICH among people taking both lecanemab and an anticoagulant, we developed a microsimulation model to examine the benefits and harms of co-prescribing both medications in a population of people with atrial fibrillation experiencing early AD.

## METHODS

### Analytic Overview

We developed a microsimulation model to estimate the health and quality-of-life effects of apixaban and lecanemab for people with early AD and atrial fibrillation (AF). AF increases the risk of ischemic stroke; anticoagulants reduce the risk and severity of ischemic stroke but increase the risk and severity of ICH. Guidelines recommend anticoagulant use in AF when the untreated risk of stroke exceeds 2% per year (15,16). Lecanemab slows the rate of cognitive decline but increases the risk of ICH. When anticoagulation and lecanemab are used together, ICH risk appears to increase further. We compared four treatment strategies: apixaban alone (*APIX*), apixaban and lecanemab (*APIX/LEC*), lecanemab alone (*LEC*), and neither. Simulated persons experience risks of ischemic stroke, ICH, cognitive decline, and death determined by past medical history and use of lecanemab and anticoagulants. Because ICH risk increases substantially with age, we modeled age-stratified outcomes, including ICH, ischemic stroke, cognitive decline, and quality-adjusted life months (QALMs) over 18 months. We performed deterministic sensitivity analyses to assess the impact of data uncertainty on model outcomes. The Mass General Brigham Review Board provided ethical approval for this study (2022P003396).

### Model Structure

Each month, simulated persons with early AD and AF experience cognitive decline and risk of ischemic stroke, ICH, or background death. The monthly risk of ischemic stroke depends on each person’s risk as measured by the CHA_2_DS_2_-VASc score and anticoagulation status (17–19). The monthly risk of ICH depends on anticoagulation use, lecanemab use, age, sex, prior ICH, prior bleeding, prior stroke, prior myocardial infarction, ischemic heart disease, and hypertension (17,18). After an ischemic stroke or ICH event, the person either dies or recovers to various levels of disability. The severity of ICH and ischemic stroke is dependent on anticoagulation status (20–22). The rate of cognitive decline is slowed by lecanemab use. Age- and sex-stratified background mortality rates increase with a history of stroke or ICH and increase with dementia severity. The model schematic can be found in **Supplemental Figure 1**.

### Input Parameters

Input parameters are listed in **Table 1** and in the **Supplemental Methods**. At model start, individuals are assigned a specific covariate profile from a cohort of the Health and Retirement Study (HRS) aged 65-90 with cognitive impairment and AF. We used the 2018-19 interview cycle of the HRS (n=262), which was scaled proportionally to represent the 100,000 simulated individuals (23). Characteristics obtained from the HRS and used in the model include: age, sex, history of stroke, bleeding, hypertension, myocardial infarction, heart failure, and diabetes (**Supplemental Table 1**). We used a validated algorithm to categorize HRS participants’ cognitive status as normal, MCI, or dementia (24). We mapped these categories to the Clinical Dementia Rating Scale Sum of Boxes (CDR-SB) score by assigning individuals to the midpoint CDR-SB score for a given category (i.e., MCI CDR-SB score 2.25; dementia CDR-SB score 6.75). At model start, 52% are female, 66% have MCI and 34% have dementia, and the mean CHA_2_DS_2_-VASc score is 4.9 (range, 2-9).

**Table 1:** Model Input Parameters to Compare 4 Strategies for Older Adults with Early AD and Atrial Fibrillation using Apixaban (*APIX*), Apixaban and Lecanemab (*APIX/LEC*), Lecanemab (*LEC*), or Neither

| Parameter | Value | Reference |
| --- | --- | --- |
| Cohort characteristics |  |  |
| Age, n (%) |  |  |
| 65-74 years | 33 (12.6%) | Derived from (23) |
| 75-84 years | 131 (50.0%) |  |
| 85+ years | 98 (37.4%) |  |
| Women, n (%) |  |  |
| 65-74 years | 20 (61) | Derived from (23) |
| 75-84 years | 60 (46) |  |
| 85+ years | 57 (58) |  |
| CHA<sub>2</sub>DS<sub>2</sub>-VASc score, mean (SD) |  |  |
| 65-74 years | 4.2 (1.4) | Derived from (17,18,23) |
| 75-84 years | 5.0 (1.7) |  |
| 85+ years | 4.9 (1.8) |  |
| Baseline CDR-SB, mean (SD) |  |  |
| 65-74 years | 3.7 (2.0) | Derived from (23,24,30) |
| 75-84 years | 3.5 (1.9) |  |
| 85+ years | 3.9 (2.1) |  |
| Intracranial hemorrhage (ICH) |  |  |
| ICH rate per 1,000 person-years |  |  |
| 65-74 years | 1.2 | (25) |
| 75-84 years | 4.3 |  |
| 85-90 years | 7.9 |  |
| Relative treatment effect of anticoagulants | 1.84 | Derived from (19) |
| Relative treatment effect of lecanemab | 2.02 | Derived from (8) |
| Relative treatment effect of anticoagulants and lecanemab* | 9.92 |  |
| Relative interaction between anticoagulants and lecanemab | 2.67 | Derived from (8,19) |
| Ischemic stroke** |  |  |
| Baseline ischemic stroke rate without anticoagulants per 1,000 person-years (specific values based on CHA<sub>2</sub>DS<sub>2</sub>-VASc score) | 22.0-122.3 | (18) |
| Relative treatment effect of apixaban alone | 0.230 | Derived from (50,51) |
| Relative treatment effect of lecanemab alone | 1.000 | Assumption |
| Cognitive decline (defined by CDR-SB Score) |  |  |
| Baseline rate of cognitive decline (increase in CDR-SB score per month) | 0.092 | (1) |
| Absolute treatment effect of lecanemab on CDR-SB score per month | -0.025 |  |
| Mortality |  |  |
| Background mortality (among people without dementia) | Age-, sex-stratified | Derived from (52,53) |
| Hazard among people with mild cognitive impairment | 1.48 | (33) |
| Hazard among people with dementia | 2.84 | (33) |
| Hazard following intracranial hemorrhage | 2.19 | (34) |
| Hazard following ischemic stroke | 1.34 | (20,54) |
| Quality of life |  |  |
| Quality of life with cognitive impairment |  |  |
| Mild cognitive impairment (CDR-SB 0.5-4.0) | 0.80 | (30,36) |
| Mild dementia (CDR-SB 4.0-9.0) | 0.74 |  |
| Moderate dementia (CDR-SB 9.0-15.5) | 0.59 |  |
| Quality of life following ICH or ischemic stroke |  |  |
| Full Recovery (GOS 5) | 0.93 | (27,55) |
| Mild impairment (GOS 4) | 0.76 |  |
| Severe impairment (GOS 3) | 0.50 |  |

\*The model base case uses a 1.84-fold increase in the risk of ICH due to anticoagulants, 2.02-fold higher due to lecanemab without anticoagulation (trial-reported) and 9.92-fold higher for people on lecanemab and anticoagulation (trial-reported). Based on these, we calculate a relative lecanemab-anticoagulant interaction on ICH of 2.67-fold.
\*\*Stratified by CHA2DS2-VASc Score.
Early AD is mild cognitive impairment or mild dementia due to Alzheimer's disease.

#### Ischemic Stroke and Intracranial Hemorrhage

The baseline rate of ischemic stroke is 22-122 events/1,000 person-years, depending on CHA_2_DS_2_-VASc score, without anticoagulants or lecanemab use (**Table 1**) (17,18). The CHA_2_DS_2_-VASc score uses age, sex, and history of heart failure, hypertension, stroke, diabetes, and vascular disease to predict ischemic stroke risk among people with AF (17). The baseline rate of ICH without anticoagulants or lecanemab is 1.2-7.9 events/1,000 person-years, depending on age and comorbidity history (25). With anticoagulants, ICH risk increases 1.84-fold (19). Based on the Clarity AD trial, the risk of ICH is 2.02-fold higher among people on lecanemab without anticoagulants and 9.92-fold higher on lecanemab and anticoagulants (1,9). Using these values, we calculated a lecanemab-anticoagulant interaction on ICH of 2.67-fold. Following ischemic stroke or ICH, individuals recover one of four health states measured by the Glasgow Outcomes Scale (GOS). We estimated transition probabilities of ischemic stroke and ICH events to health states from published data (**Supplemental Table 2**) (20–22,26,27).

#### Cognitive Decline

To mirror the Clarity AD trial, we modeled cognitive decline using the CDR-SB, which is a clinician-rated scale with a range of 0 to 18. In the model, people experience cognitive decline at the rate observed in the placebo arm of the Clarity AD trial—a rise in CDR-SB score of 0.092/month so that the mean progression from MCI to mild dementia occurs in 38 months (1). Lecanemab use slows the rise in CDR-SB score by 0.025/month (i.e., CDR-SB worsening slows from 0.092/month to 0.067/month) (1).

#### Quality of Life

We mapped model-estimated CDR-SB scores to clinical categories of cognitive impairment and then to quality of life: normal cognition (CDR-SB <0.5; QoL 1.00), MCI (CDR-SB 0.5-4.0; QoL 0.80), mild dementia (CDR-SB 4.0-9.0; QoL 0.74), moderate dementia (CDR-SB 9.0-15.5; QoL 0.59), and severe dementia (CDR-SB 15.5-18.0; QoL 0.36) (28–30). Quality of life estimates were linearly smoothed across clinical categories of cognition to avoid large step changes when an individual progresses to a more severe cognitive state (**Supplemental Figure 2**). After a stroke or ICH event, people recover to a GOS level with associated quality of life between 0.93 (full recovery) and 0 (death) (31). We multiplied quality of life estimates when more than one health state was present.

#### Mortality

We first determined baseline age- and sex-stratified mortality rates among people without dementia (32). Then, based on the degree of cognitive decline, we applied an excess mortality hazard associated with cognitive impairment: MCI (HR 1.48) and dementia (HR 2.84) (33). This hazard was smoothed linearly across clinical categories of cognition such that there was no abrupt discontinuity in hazard if an individual progressed to a more severe cognitive state (**Supplemental Figure 3**). Nonfatal strokes and intracranial hemorrhages increased the mortality hazard in subsequent periods (HR 1.34 and HR 2.19, respectively) (20,34).

### Sensitivity and Scenario Analyses

We performed one-way sensitivity analyses to examine the effect of parameter uncertainty on model outcomes, as well as multiway sensitivity analyses to assess the implications of varying multiple influential parameters simultaneously (35). The range over which parameters were tested in sensitivity analyses reflected their uncertainty, particularly trial-derived estimates with small numbers (**Supplemental Table 3**). Given different approaches to estimating the quality of life associated with cognitive impairment, we conducted a scenario analysis using quality-of-life estimates that are community-standard and patient-reported, in contrast to the caregiver proxy measure used in the base case (**Supplemental Table 4**) (36).

## RESULTS

### Base case

Age-stratified model outcomes are shown in **Table 2**. For people aged 65-74 years, apixaban alone (*APIX*), apixaban plus lecanemab (*APIX/LEC*), and lecanemab alone *(LEC)* would each result in 13.2 QALMs over 18 months but with substantial differences in clinical events. Compared to *APIX*, *APIX/LEC* would result in more ICHs per 100,000 persons (1,990 vs. 400), more all-cause mortality (5,820 vs. 5,140 deaths), and slower cognitive decline (mean change in CDR-SB, 1.11 vs. 1.53) over 18 months. Compared to *APIX*, *LEC* would result in a similar number of ICHs (420 vs. 400), more ischemic strokes (8,580 vs. 1,900), more all-cause mortality (5,770 vs. 5,140 deaths), and slower cognitive decline (mean change in CDR-SB 1.11 vs. 1.53).

**Table 2:** Projected Clinical Impact of Strategies Using Apixaban (*APIX*), Apixaban and Lecanemab (*APIX/LEC*), Lecanemab (*LEC*), or Neither for 100,000 Simulated Adults with Early AD and Atrial Fibrillation over 18 months

|  | **APIX** | **APIX/LEC** | **LEC** | **Neither** |
| --- | --- | --- | --- | --- |
| **Number of Intracranial Hemorrhage (ICH) Events** |  |  |  |  |
| 65-74 years | 400 | 1,990 | 420 | 220 |
| 75-84 years | 1,260 | 7,090 | 1,400 | 750 |
| 85-90 years | 2,260 | 12,590 | 2,570 | 1,190 |
| **Mean Change in Clinical Dementia Rating Scale Sum of Boxes (CDR-SB) Score\*** |  |  |  |  |
| 65-74 years | 1.53 | 1.11 | 1.11 | 1.52 |
| 75-84 years | 1.47 | 1.05 | 1.06 | 1.46 |
| 85-90 years | 1.38 | 0.98 | 1.01 | 1.38 |
| **Number of Ischemic Stroke Events** |  |  |  |  |
| 65-74 years | 1,900 | 1,860 | 8,580 | 8,300 |
| 75-84 years | 2,250 | 2,220 | 9,690 | 9,670 |
| 85-90 years | 2,050 | 1,980 | 9,050 | 9,100 |
| **Number of All-Cause Deaths** |  |  |  |  |
| 65-74 years | 5,140 | 5,820 | 5,770 | 5,900 |
| 75-84 years | 12,660 | 15,860 | 13,130 | 13,380 |
| 85-90 years | 22,330 | 27,350 | 22,160 | 22,460 |
| **Mean Quality-Adjusted Life Months** |  |  |  |  |
| 65-74 years | 13.2 | 13.2 | 13.2 | 13.1 |
| 75-84 years | 12.8 | 12.6 | 12.7 | 12.6 |
| 85-90 years | 12.0 | 11.6 | 12.0 | 11.9 |

\*Clinical Dementia Rating scale Sum of Boxes (CDR-SB) scores range from 0 to 18, where a higher score indicates more cognitive impairment.

For people aged 75-84 years, *APIX/LEC* would reduce quality-adjusted life months (mean 12.6 vs. 12.8 QALMs), increase ICH events (7,090 vs. 1,260), slow cognitive decline (mean change in CDR-SB, 1.05 vs. 1.47), and increase all-cause mortality (15,860 vs. 12,660 deaths) per 100,000 people, compared to *APIX*.

*APIX/LEC* would reduce QALMs (mean 11.6 vs. 12.0) for people aged 85-90 years, increase ICH events (12,590 vs. 2,260), slow cognitive decline (mean change in CDR-SB, 0.98 vs. 1.38), and increase all-cause mortality (27,350 vs. 22,330 deaths) per 100,000 people, compared to *APIX*.

### One-Way Sensitivity Analyses

Results from the one-way sensitivity analysis show that lecanemab’s effect on cognitive decline is the most sensitive parameter in the direction of increasing preference for *APIX/LEC* for people 65 to 74 years old (**Figure 1**). The preference for *APIX/LEC* increases when the effect of lecanemab on slowing CDR-SB is greater than 0.45 over 18 months. If the effect of lecanemab on CDR-SB is 1.66 points, meaning it halts cognitive decline for 18 months, *APIX/LEC* would produce a gain of 0.2 QALMs compared to *APIX*, despite 1590 excess ICHs. Model outcomes are not sensitive to the effect of cognitive impairment on mortality.

**Figure 1:**
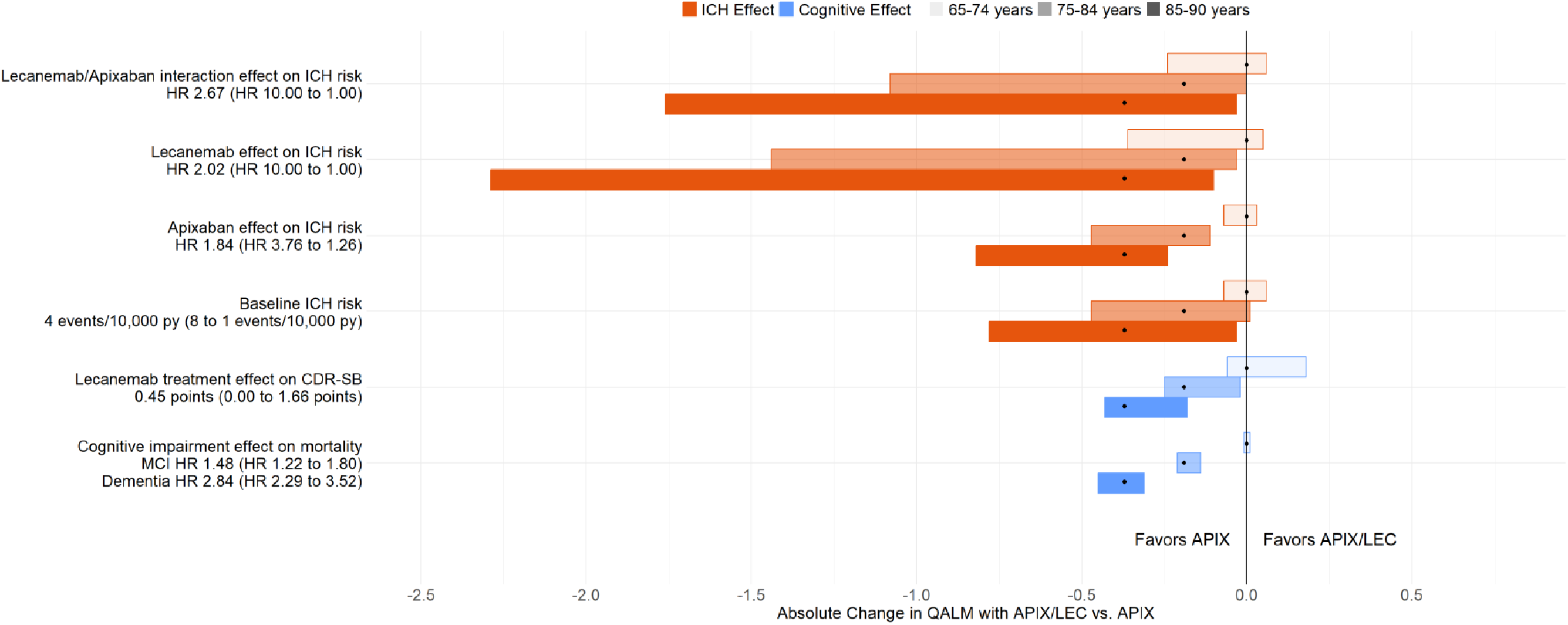
One-way Sensitivity Analyses Comparing Model-projected QALMs for Apixaban alone (APIX) vs. Apixaban and Lecanemab (APIX/LEC), Stratified by Age for Older Adults with Early AD and Atrial Fibrillation. <u>Legend</u> This tornado diagram shows the absolute difference in QALMs, comparing the strategies of *APIX* and *APIX/LEC* for people with atrial fibrillation and early AD (mild cognitive impairment or mild dementia due to Alzheimer’s Disease). Input parameters are displayed along the vertical axis on the left, followed by the corresponding unit, base case value, and the range in parentheses. The input value resulting in favoring *APIX* is listed before the hyphen, and the input value that results in favoring *APIX/LEC* is listed after the hyphen. The bold vertical line denotes no difference in QALMs between the strategies. Each black dot represents the base case for each age strata, and the width of the horizontal bar represents the range of QALMs over the tested range of input values for each model parameter, holding all other parameters constant. We detail the rationale for parameter ranges in Supplemental Table 2. ICH – intracranial hemorrhage; py – person-year; CDR-SB – Clinical Dementia Rating scale Sum of Boxes; QALM – quality-adjusted life month; MCI – mild cognitive impairment due to Alzheimer’s disease

For people 65 to 74 years, clinical benefit was sensitive to four determinants of ICH risk. *APIX/LEC* would be preferred to *APIX* if: there is no interaction between lecanemab and apixaban (+0.1 QALM); the baseline risk of ICH is 1 in 10,000 person-years, (+0.1 QALM); lecanemab has no effect on ICH risk (+0.1 QALM); apixaban only increases the risk of ICH 1.26-fold (<0.1 QALM). For people aged 75-90 years, using plausible parameter estimates we found no scenario in which *APIX/LEC* would result in more clinical benefit when compared to *APIX*. One-way sensitivity analyses for ICH, CDR-SB, and overall mortality are displayed in **Supplemental Figure 4**.

### Multiway Sensitivity Analyses

In multiway sensitivity analyses, we compared *APIX* to *APIX/LEC* among people 65-74 years and varied the lecanemab-apixaban interaction against the lecanemab effect on ICH and the lecanemab effect on CDR-SB (**Figure 2A** and **2B**, respectively). Among people 65-74 years old, *APIX* alone would be the preferred strategy when lecanemab has no effect on CDR-SB (i.e., 0 point change over 18 months), at higher estimates of a lecanemab-apixaban interaction (≥4.89-fold), or a combination of the two (e.g., ≤0.22 CDR-SB points over 18 months and ≥3.78-fold lecanemab-apixaban interaction) (**Figure 2A**; open, solid, and hash square, respectively). *APIX/LEC* would be the preferred strategy if lecanemab has greater efficacy on CDR-SB (≥0.79 points over 18 months), lecanemab and apixaban do not interact (i.e., interaction effect 1.0), or a combination of both factors (e.g., ≥0.56 CDR-SB points over 18 months and ≤1.56-fold lecanemab-apixaban interaction) (**Figure 2A**; open, solid, and hash diamond, respectively).

**Figure 2:**
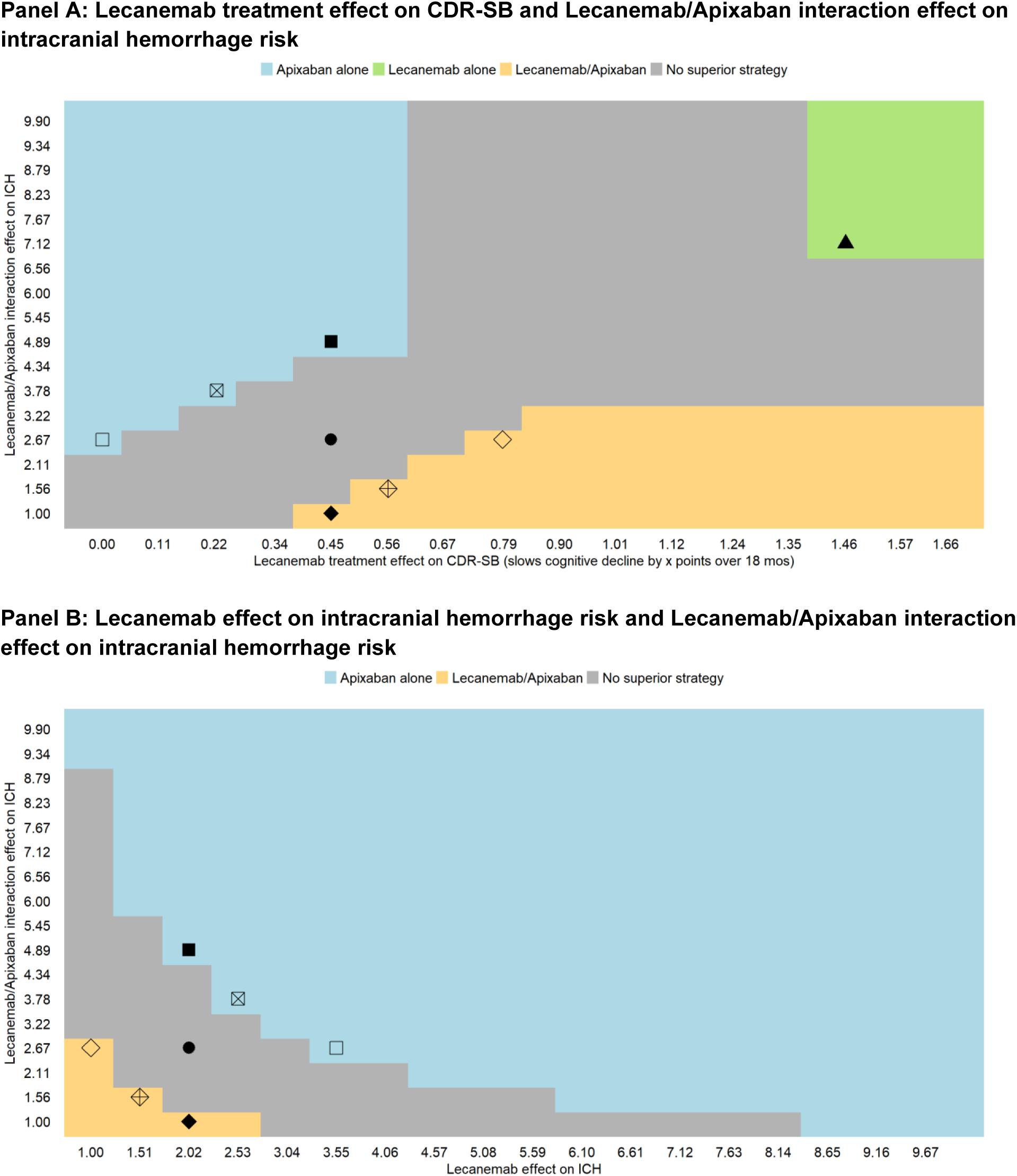
Multiway Sensitivity Analyses Comparing Strategies Using Apixaban (*APIX*), Apixaban and Lecanemab (*APIX/LEC*), Lecanemab (*LEC*), or Neither for people aged 65-74 years with early AD and Atrial Fibrillation. <u>Legend</u> Multiway sensitivity analyses comparing the quality-adjusted life months (QALMs) of apixaban alone (*APIX*), apixaban and lecanemab (*APIX/LEC*), and lecanemab alone (*LEC*). In both panels, the lecanemab-apixaban interaction is varied against lecanemab’s effect on CDR-SB (panel A) and lecanemab’s effect on ICH (panel B). The base case is marked by a solid dot. The squares represent values where *APIX* is preferred, diamonds identify values where *APIX/LEC* are preferred, and triangle where *LEC* is preferred. The solid fill represents the base-case value of the parameter on the x-axis whereas the open fill represents the base-case value of the parameter on the y-axis. The hash fill represents values where neither parameter is at its base value. CDR-SB scores range from 0 to 18; a higher score indicates more cognitive impairment. In panels A-C, the lecanemab effect on CDR-SB is an absolute difference between untreated and treated with lecanemab; thus, higher values represent a greater effect of lecanemab. We limited precision to 0.1 quality-adjusted life months (i.e., 3 quality-adjusted life days); when the difference in QALMs of two strategies is less than 0.1, we display that region in gray. CDR-SB – Clinical Dementia Rating scale Sum of Boxes; QALM – quality-adjusted life month.

When the effect of lecanemab on ICH increases (≥3.55-fold), or the lecanemab-apixaban interaction increases (≥4.89-fold), or a combination of both (e.g., ≥2.53-fold effect of lecanemab on ICH and ≥3.78-fold lecanemab-apixaban interaction), *APIX* would be the preferred strategy (**Figure 2B**; open, solid, hash square, respectively). Conversely, when there is no effect of lecanemab on ICH, no lecanemab-apixaban interaction on ICH, or both are lower (e.g., ≤1.51-fold effect of lecanemab on ICH and <1.56-fold lecanemab-apixaban interaction), *APIX/LEC* would be the preferred strategy (**Figure 2B**; open, solid, hash diamond, respectively). Additional multiway sensitivity analyses are displayed in **Supplemental Figure 5 A-D**.

### Quality of life associated with cognitive impairment: scenario analysis

Different quality-of-life estimates for cognitive impairment affect projected QALMs (**Supplemental Table 5**). When using patient-rated quality-of-life estimates, all projected QALMs increase given the higher quality-of-life estimates using this method. However, relative differences between strategies are similar, as well as age-related differences; thus, the primary results are robust to the method used to estimate the quality of life associated with cognitive impairment.

## DISCUSSION

Atrial fibrillation co-occurs in 12% of the 6.5 million people with AD in the United States, and the tradeoffs between the use of anticoagulation and lecanemab are uncertain (12–14). These model-based results support anticoagulants without lecanemab as the preferred strategy for people with early AD and atrial fibrillation and contribute evidence regarding the tradeoffs of the concurrent use of anticoagulants and lecanemab. While adding lecanemab to anticoagulants would slow cognitive decline, it would also substantively increase the number of intracranial hemorrhages and all-cause mortality. These results are consistent with the recent Appropriate Use Recommendations guidance to avoid co-prescribing anticoagulants and lecanemab and support Medicare’s coverage decision, which requires prospective comparative studies to define the benefits and harms of lecanemab more precisely when co-prescribed with anticoagulants (9,10).

While these results support a strategy of anticoagulation without the addition of lecanemab for people with co-morbid atrial fibrillation and AD, there is important variation in outcomes when stratified by age. To integrate the small improvement in cognition and an increase in the risk of rare but often devastating ICHs, we used quality-adjusted life months to quantify clinical outcomes and considered changes of less than 0.1 QALM to be too small to be clinically relevant. For people aged 65 to 74, adding lecanemab to anticoagulants would result in modest population-wide benefit due to slower cognitive decline that would be offset by an increase in rare, but devastating ICH events. These limited health effects should be interpreted in the context of the cost of prescribing lecanemab—the drug costs $26,500 annually, with an additional $82,500 in annual ancillary costs related to diagnosis, administration, and safety monitoring (37). In contrast, for people older than 75 years anticoagulation alone is the preferred strategy even ignoring cost. While the addition of lecanemab to anticoagulation would slow cognitive decline, these benefits would be outweighed by the harms resulting from the marked increase in intracranial hemorrhage events, driven by the higher baseline risk of ICH in this age group.

The uncertainty surrounding the clinical relevance of lecanemab’s effect on cognitive decline is vital when interpreting the study results. While Clarity AD showed a statistically significant reduction in the rate of cognitive decline with lecanemab, as measured by the change in CDR-SB score, some argue that the absolute difference is small and may not be clinically meaningful (5–7). The model used in this analysis assumes that people derive mortality and quality of life benefits from even the most marginal slowing of cognitive decline. However, if a clinical benefit is not appreciable over the 18-month period tested in the trial, then these results may overstate the expected benefit of any strategies that use lecanemab.

These findings have at least three implications for current practice and future work. First, we identify clear targets for post-approval comparative effectiveness studies. As with any new therapeutic, there is uncertainty about lecanemab’s efficacy and side effects. For people aged 65 to 74 years, the clinical benefit, in QALMs, is sensitive to three drug-specific inputs—the effect of lecanemab on ICH, the lecanemab-anticoagulant interaction, and the effect of lecanemab on cognitive decline. The Clarity AD trial, the phase 3 trial testing lecanemab, was likely underpowered to estimate the lecanemab-anticoagulant interaction and to determine if this interaction varies by anticoagulant type (1,38). Additionally, it is unclear if the effect of lecanemab is heterogeneous—that is, if lecanemab slows cognitive decline to a greater or lesser extent in some groups. More precise estimates in post-trial observational studies could help clarify the preferred strategy for people aged 65 to 74 years.

Second, these results highlight the importance of person-specific ICH risk factors in the decision to use lecanemab and anticoagulants together. Specifically, adding lecanemab to anticoagulants may produce an overall clinical benefit when the risk of ICH is lower than average, as it is in younger age groups. These model-based results show the potential benefits of considering baseline ICH risk prior to initiating lecanemab. The results are concordant with the FDA’s black box warning and expert guidance to determine APOE4 status, a known risk factor for ICH in general, and in post-publication analyses of Clarity AD (11,38). Additional screening strategies could be beneficial, such as the use of more sensitive MR imaging to identify cerebral microhemorrhage and cortical superficial siderosis, which are markers of concurrent cerebral amyloid angiopathy pathology and predictors of future ICH (39–41).

Third, these results are germane for all older adults being evaluated for lecanemab since many older adults develop new anticoagulant indications over time. For example, among adults aged 65 years and older, the incidence of atrial fibrillation is 23.7 per 1,000 person-years, and the incidence of venous thromboembolism is 4.1 to 9.0 per 1,000 person-years (42,43). The diagnosis of such conditions during a lecanemab treatment course poses a challenge (44). These study results can inform patient, care partner, and clinician decision-making by providing data on the risks and benefits of concurrent therapy.

These results are relevant to the care of people with early AD despite the recent FDA approval of donanemab. TRAILBLAZER-ALZ 2, the phase 3 trial comparing donanemab to placebo for early AD, demonstrated that donanemab was similarly effective at slowing cognitive decline, and unpublished data from the same trial indicated that donanemab may not interact with anticoagulants to increase ICH risk, though the trial was not designed to detect such an interaction (45,46). Nevertheless, the possible advantages of donanemab must be balanced against the higher reported rates of amyloid-related imaging abnormalities (ARIA) with edema or effusion (ARIA-E) and ARIA with hemosiderin deposits (ARIA-H), the principal adverse effects of anti-amyloid antibody therapy. While lecanemab and donanemab both increase the risk of ARIA, TRAILBLAZER-ALZ 2 reported an absolute risk increase of overall ARIA-E and ARIA-H rates attributable to donanemab that was double what Clarity AD reported for lecanemab (1,46). As such, it is not clear if donanemab will be preferred over lecanemab in people with early AD regardless of concurrent anticoagulant use. Consideration of the risks and benefits of each pharmacologic intervention and further data collection to estimate ICH incidence more precisely will be important next steps in determining the preferred strategy for people with early AD.

The study findings have important limitations. First, this model does not account for decrements in quality of life attributable to the treatment administration and logistics, self-monitoring, or ARIA without ICH. The effect of ARIA and monitoring for ARIA on quality of life was not reported by the clinical trial, and no post-approval data exist to our knowledge. Thus, to the degree ARIA without ICH affects quality of life, these results could overestimate the true benefit of lecanemab. Second, this study used apixaban for thromboprophylaxis in atrial fibrillation. There are no data on a potentially heterogeneous interaction between lecanemab and anticoagulants by anticoagulant type, and this model did not test non-anticoagulant thromboprophylaxis strategies, such as left atrial appendage occlusion. The study also did not examine other indications for anticoagulation, e.g., valve replacement. Third, individual patients could weigh the significance or quality-of-life implications of each outcome differently from the population-level estimates used in the study. Additionally, there is uncertainty related to the model inputs which are derived from a variety of sources. Finally, this simulation modeled the effect of lecanemab on CDR-SB over 18 months to reflect the evidence produced by Clarity AD. While a longer treatment course has been proposed by some, including the manufacturer of lecanemab, there are limited data on the treatment effect beyond 18 months (47).

In sum, in this model-based analysis, we find that apixaban without lecanemab is the preferred strategy for older adults aged 75+ years with early AD and atrial fibrillation. Although adding lecanemab could slow cognitive decline, it would likely result in a greater number of ICHs and deaths among people who are concurrently anticoagulated. Notably, there is greater equipoise for people aged 65 to 74 years, where anticoagulation with lecanemab would risk more deaths due to ICHs but yield some quality of life benefits from the slower progression of cognitive decline. These results should inform current guidance on the concurrent prescription of anticoagulants and lecanemab and highlight that improved estimates of the lecanemab-anticoagulant interaction are critical to identifying the preferred strategy for people aged 65-74 years with comorbid atrial fibrillation and early AD.

## Data Availability

Data are available after consent/use agreements executed from the Health and Retirement Study (https://hrsdata.isr.umich.edu/) study coordinators. Input parameters for simulation model derived from sources cited in manuscript (Table 1, supplemental Tables 2, 3).

## Supplemental Methods – Model Structure and Parameterization

### 1. Model Overview

This is an individual-level microsimulation with a monthly time step. The time horizon mirrors the Clarity AD trial, which tested the effect of lecanemab over 18 months. At model start, all simulated individuals have cognitive impairment and atrial fibrillation. Each month, they experience cognitive decline, a risk of ischemic stroke (IS), a risk of intracranial hemorrhage (ICH), and a risk of death. **Supplemental Figure 1** displays the model schematic.

After an IS or ICH event, a simulated person can transition to one of the following health states related to physical function—full recovery, minor disability, major disability, or death. In parallel, simulated individuals continue to experience cognitive decline. Health states related to cognition include: mild cognitive impairment, mild dementia, or moderate dementia.

### 2. Simulated population: data sources

#### a. Overview

In Section 2, we detail the data elements from the Medicare-linked Health and Retirement Study. In Section 3, we describe how we use these data to populate the model.

#### b. The Health and Retirement Study (HRS)

The HRS is a nationally representative, longitudinal study of older Americans (23). Participants are age 50 and older and are interviewed every two years to measure changes in disability, health, and wealth as they transition from work to retirement. The study interviews participants related to four broad areas of aging: income and wealth; health, function, and cognition; work and retirement; and family connections. If a participant was unable to complete an interview because of physical or cognitive impairment, the interview was conducted with a proxy, usually a family member. For the participants who provided consent, the HRS also provides linkages to administrative data, including Medicare insurance claims. Through sampling methodology, the HRS is nationally representative of the community-dwelling and nursing home population of older Americans (56). Population estimates of mortality, education, and nursing home residence from the HRS match estimates from the US Census (57).

#### c. Study population identification

To identify the data that will be used to populate the model, we considered HRS participants who completed the 2018-19 interview, were 65-90 years or older at the time of the interview, agreed to Medicare claims linkage, and were continuously enrolled in Medicare fee-for-service (Part A and B) in the 12 months before their 2018 interview. We included participants who had one inpatient or two outpatient claims for atrial fibrillation. All observations had a CHA_2_DS_2_-VASc score corresponding to an ischemic stroke risk ≥ 2% / year. In prior studies, we have also used this approach to identify people in the HRS with AF (48,49). We only included participants with cognitive impairment (MCI or dementia), described in greater detail below.

#### d. Cognitive impairment classification

To assess cognition, HRS administers a validated 27-question neuropsychological battery to participants or an informant questionnaire to proxies when participants cannot be interviewed (24). We used the Langa Weir model which uses the responses to these 27 items to probabilistically categorize individuals as normal, cognitive impairment not dementia, or dementia which correlates with clinical diagnoses of no impairment, MCI, and dementia, respectively. The Langa Weir model was validated against a neuropsychiatric diagnosis (58).

#### e. Mapping cognitive impairment to CDR-SB

Because the HRS does not measure the CDR-SB, we translated cognitive categories to corresponding CDR-SB scores. O’Bryant et al. identified the CDR-SB score ranges that correspond to dementia staging categories (30). Relevant to this study, MCI corresponds to a CDR-SB range of 0.5 to 4.0 and mild dementia to a range of 4.0 to 9.0. We assigned each individual in the model a starting CDR-SB score that corresponded to the midpoint of the range for their cognitive category, i.e., MCI starting score of 2.25 and dementia starting score of 6.50.

#### f. Individual characteristics

Beyond atrial fibrillation and cognitive function, the following characteristics were obtained using a combination of the HRS survey and Medicare claims data: age, sex, history of stroke, bleeding, hypertension, myocardial infarction, heart failure, and diabetes.

### 3. Simulated population: cohort characteristics

At model start, individuals draw for a specific profile of covariates that corresponds to individuals in the 2018 Health and Retirement Study (HRS) with atrial fibrillation and cognitive impairment. We proportionally scaled this cohort of 262 to 100,000 simulated individuals.

### 4. Model parameterization: cognitive decline

#### a. CDR-SB

Cognitive decline is often measured in clinical trials using the Clinical Dementia Rating (CDR), a clinician-rated scale. The CDR rates individuals in 6 functional areas that are scored as 0 (no impairment), 0.5 (questionable impairment), 1 (mild dementia), 2 (moderate dementia), or 3 (severe dementia). The CDR sum of boxes (CDR-SB) sums all the scores with a range of 0 to 18. While the scale is ordinal, we modeled it as a continuous scale to simulate the monthly progression of cognitive impairment in order to account for the associated changes in mortality risk and quality of life.

#### b. Mapping incremental changes in CDR-SB to quality of life (QoL)

Studies of QoL corresponding to cognition status have measured the QoL effect given a cognitive category (i.e., MCI, dementia). To assign a change in QoL over an incremental change in CDR-SB we took the following steps. First, we assigned each category to a corresponding CDR-SB score. Then, we took the estimated QoL for that category and assigned it to the mid-point value for that CDR-SB range. We repeated this for each clinical category. We then linearly smoothed across categories to avoid large step changes when individuals progress to more severe cognitive states. This linear smoothing is displayed in **Supplemental Figure 2**.

#### c. Mapping incremental changes in CDR-SB to mortality risk

Studies of mortality risk corresponding to cognition status have measured the effect given a cognitive category (i.e., MCI, dementia). To assign a change in mortality risk over an incremental change in CDR-SB, we took the following steps. First, we assigned each category to a corresponding CDR-SB score. Then, we took the estimated mortality risk for that category and assigned it to the mid-point value for that CDR-SB range. We repeated this for each clinical category. We then linearly smoothed across categories to avoid large step changes when individuals progress to more severe cognitive states. This linear smoothing is displayed in **Supplemental Figure 3**.

### 5. Model parameterization: Intracranial hemorrhage (ICH)

#### a. ICH risk and calibration

We used the analysis by Friberg et al. to estimate an individual’s risk of ICH conditional on observed characteristics including age, sex, prior ICH, prior ischemic stroke, prior bleeding, hypertension, and prior myocardial infarction (18). We calibrated the ICH risk so that the ICH rate (off-lecanemab, off-anticoagulants) matched population estimates (25)

#### b. Outcome health states

Following an ICH event, individuals can transition into 1 of 4 health states as measured by the Glasgow Outcome Scale (death, severe disability, mild disability, and full recovery). ICH events that occur on anticoagulants are more likely to result in a more disabled health state—probabilities of transitioning to each health state on- and off-anticoagulants are displayed in **Supplemental Table 2**. (21)

### 6. Model parameterization: Ischemic stroke (IS)

#### a. Risk

We used the CHA_2_DS_2_-VASc score to estimate an individual’s risk of IS, conditional on observed characteristics including age, sex, congestive heart failure, hypertension, diabetes, prior stroke, and vascular disease (17).

#### b. Outcome health states

The most reliable estimates of recovery following ischemic stroke measure function use the modified Rankin Scale. To make the health states comparable to the post-ICH health states, we mapped the modified Rankin Scale to the Glasgow outcome scale based on Gaastra et al. (26).

| **Modified Rankin Scale** | **Glasgow Outcome Scale** |
| --- | --- |
| No symptoms (mRS 0) | Full recovery (GOS 5) |
| No significant disability despite symptoms (mRS 1) | Full recovery (GOS 5) |
| Slight disability (mRS 2) | Mild disability (GOS 4) |
| Moderate disability (mRS 3) | Severe disability (GOS 3) |
| Moderately severe disability (mRS 4) | Severe disability (GOS 3) |
| Severe disability (mRS 5) | Severe disability (GOS 3) |
| Death (mRS 6) | Death (GOS 1) |

Ischemic strokes that occur on anticoagulants are less likely to result in a disabled health state—probabilities of transitioning to each health state on- and off-anticoagulants are displayed in Supplemental Table 2 (22).

---

**Supplemental Figure 1:**
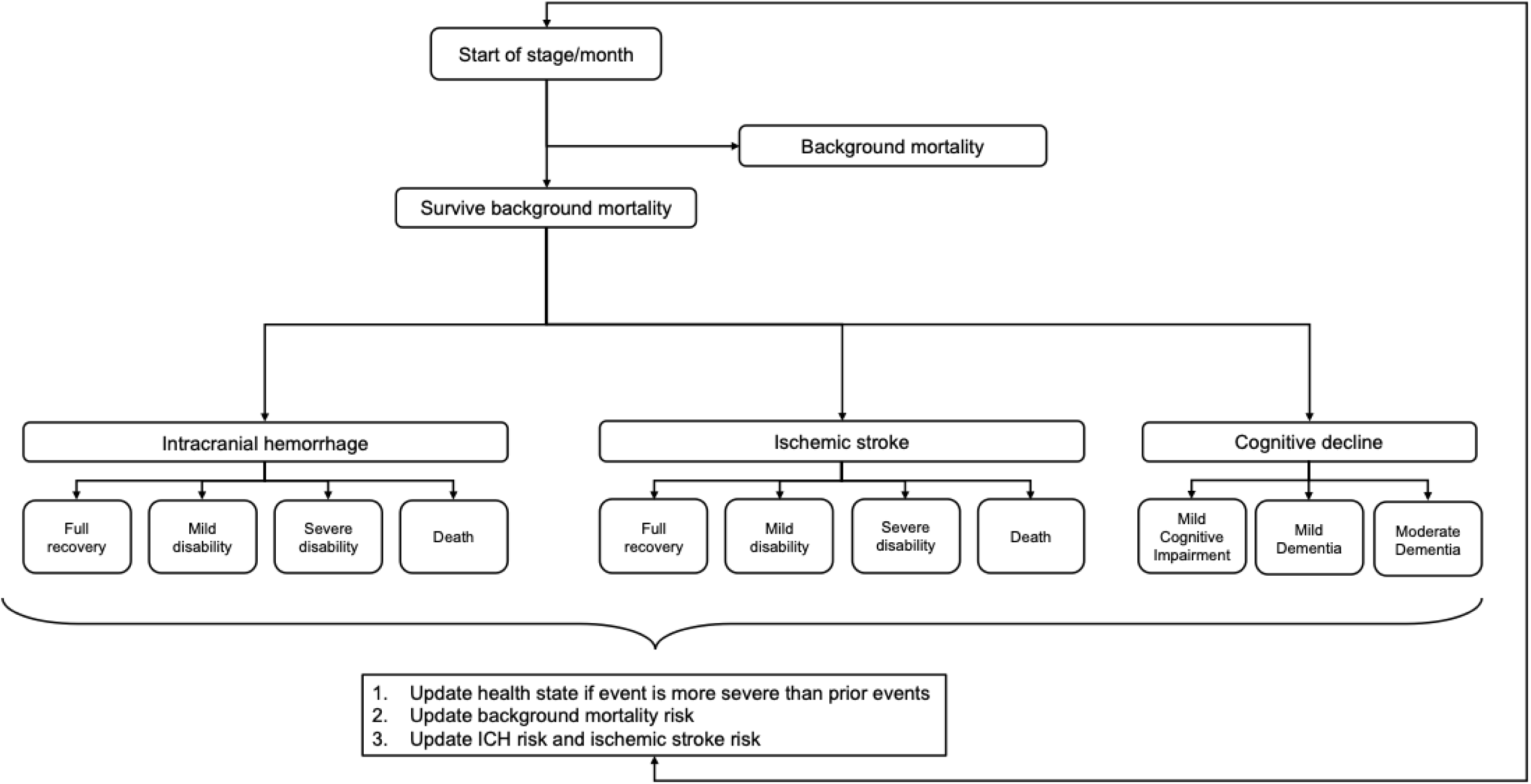
Model Schematic. <u>Legend</u> The model schematic represents the possible events and outcomes in each monthly cycle. At the beginning of each cycle, agents face a risk of death mortality from competing causes. The agents that survive then face a risk of intracranial hemorrhage and ischemic stroke, both of which can result in health states ranging from full recovery to death. All agents experience cognitive decline. At the end of each month, agents’ health state, background mortality risk, ICH risk, and ischemic stroke risk are updated.

**Supplemental Figure 2:**
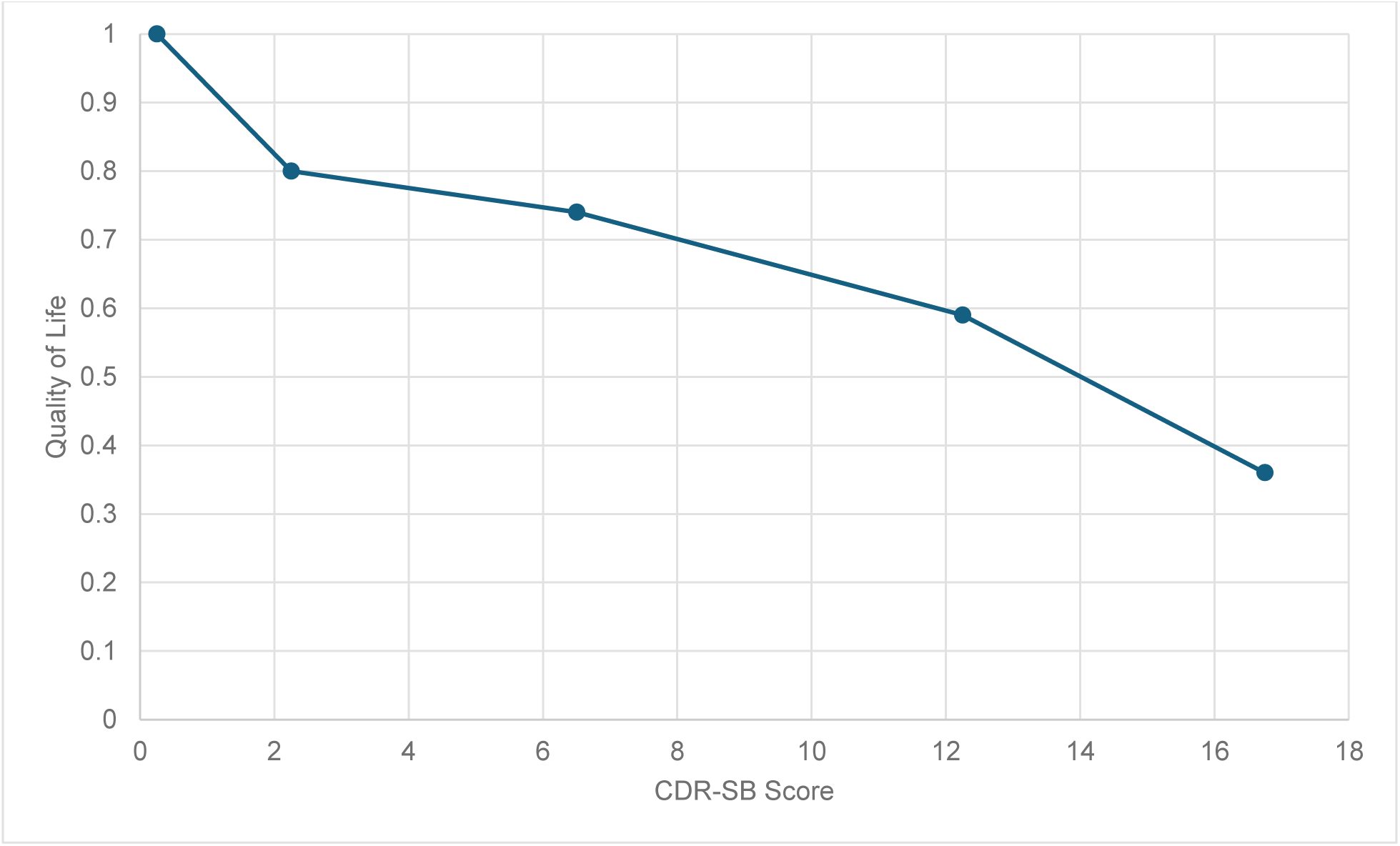
Linear smoothing of quality of life by CDR-SB score. <u>Legend</u> CDR-SB – Clinical Dementia Rating scale Sum of Boxes This plot provides a visual representation of the linear smoothing of quality of life estimates by CDR-SB score. Each cognitive category was assigned a corresponding CDR-SB score and the quality of life for that category was assigned to the mid-point value for that CDR-SB range. We linearly smoothed across categories to avoid large step changes when individuals progress to more severe cognitive states. For CDR-SB scores > 16.75, changes in quality of life were assumed to follow the same slope as in the segment immediately prior.

**Supplemental Figure 3:**
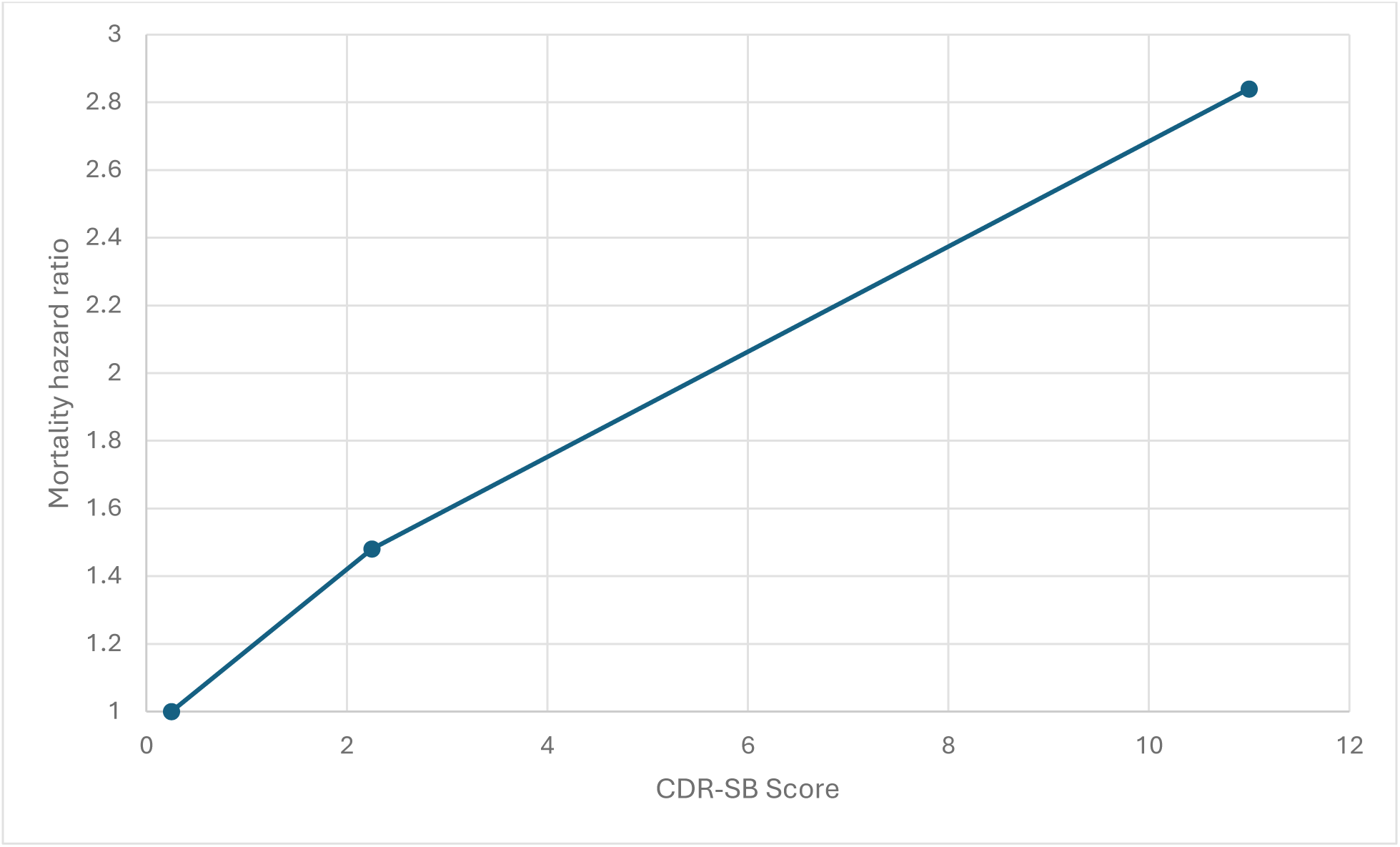
Linear smoothing of mortality hazard by CDR-SB score. <u>Legend</u> CDR-SB – Clinical Dementia Rating scale Sum of Boxes This plot provides a visual representation of the linear smoothing of the dementia-associated mortality hazard by CDR-SB score. Each cognitive category was assigned a corresponding CDR-SB score and the mortality hazard for that category was assigned to the mid-point value for that CDR-SB range. We linearly smoothed across categories to avoid large step changes when individuals progress to more severe cognitive states. For CDR-SB scores >11, changes in quality of life were assumed to follow the same slope as in the segment immediately prior.

**Supplemental Figure 4:**
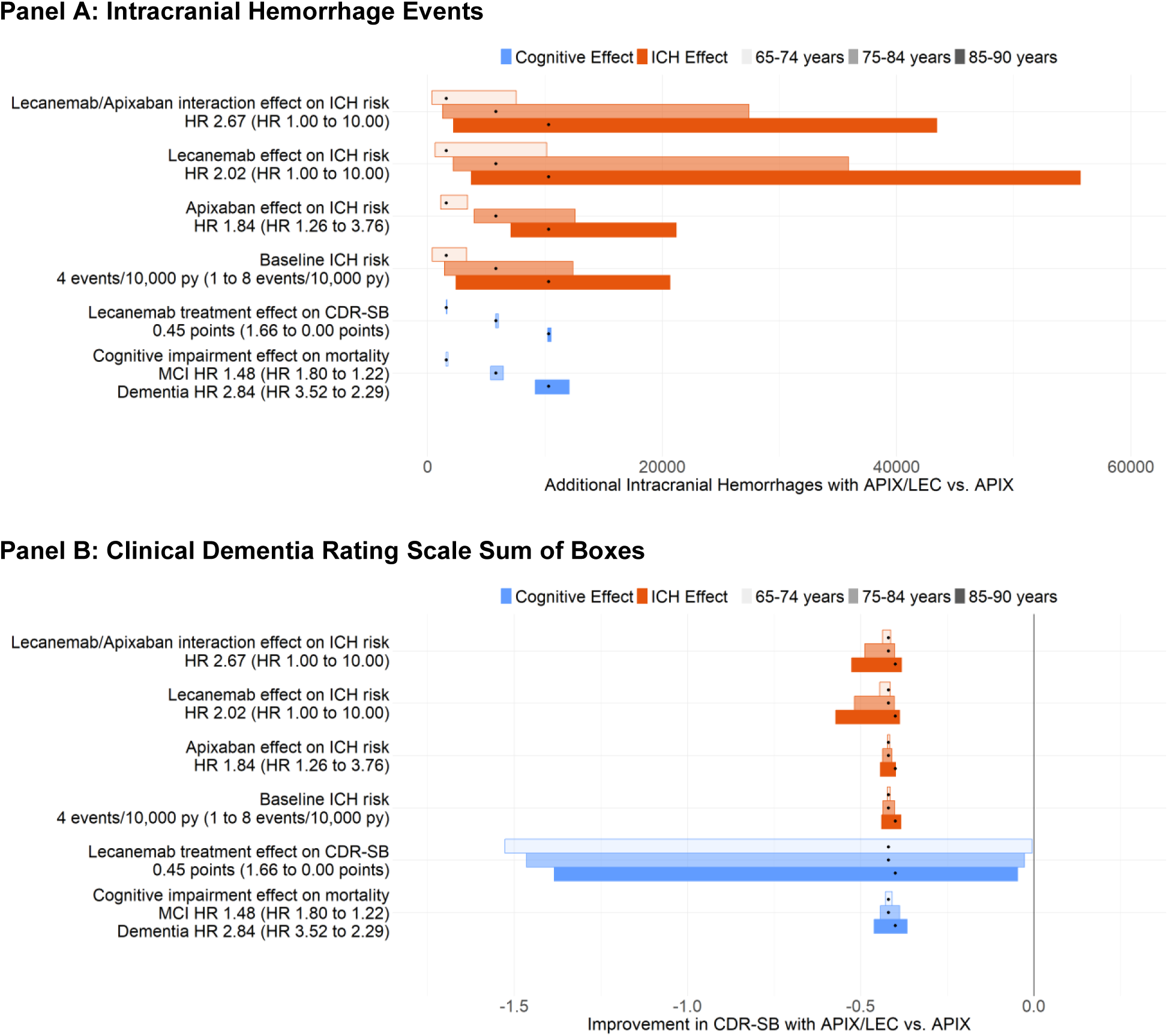
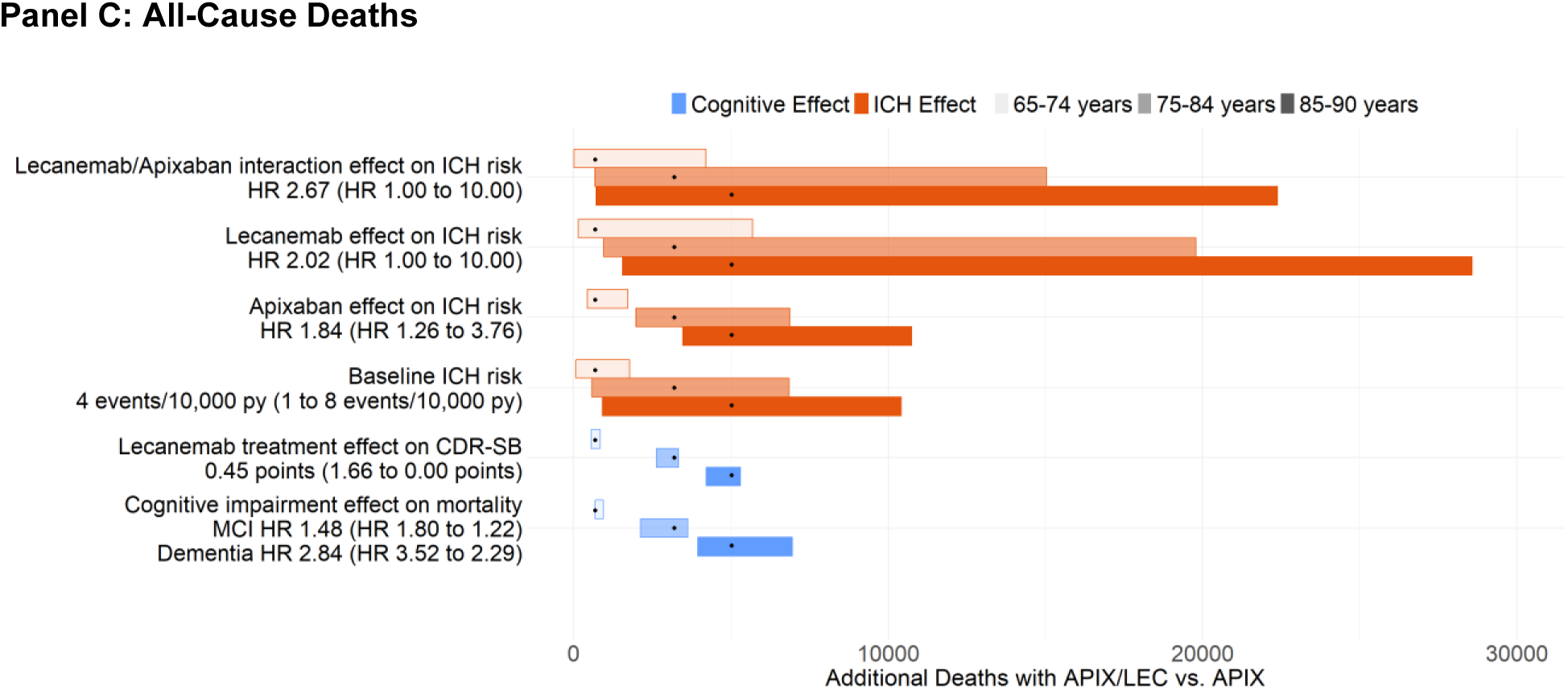
One-way Sensitivity Analyses Comparing Model-projected Health Events for Apixaban alone (*APIX*) vs. Apixaban and Lecanemab (*APIX/LEC*). Stratified by Age for Older Adults with Early AD and Atrial Fibrillation. <u>Legend</u> These tornado diagrams show the absolute difference in model-predicted intracranial hemorrhage events (Panel A), change in CDR-SB (Panel B), and all-cause deaths (Panel C), comparing the strategies of *APIX* and *APIX/LEC* for people with atrial fibrillation and early AD (mild cognitive impairment or mild dementia due to Alzheimer’s Disease). Input parameters are displayed along the vertical axis on the left, followed by the corresponding unit in parentheses, base case value, and the range in parentheses. The input value resulting in favoring *APIX/LEC* is listed before the hyphen, and the input value that results in favoring *APIX* is listed after the hyphen. The bold vertical line denotes no difference in CDR-SB between the strategies (Panel B). Each black dot represents the base case for each age strata, and the width of the horizontal bar represents the range in outcome over the tested range of input values for each model parameter, holding all other parameters constant. We detail the rationale for parameter ranges in Supplemental Table 2. ICH – intracranial hemorrhage; py – person-year; CDR-SB – Clinical Dementia Rating scale Sum of Boxes; QALM – quality-adjusted life month; MCI – mild cognitive impairment due to Alzheimer’s disease

**Supplemental Figure 5:**
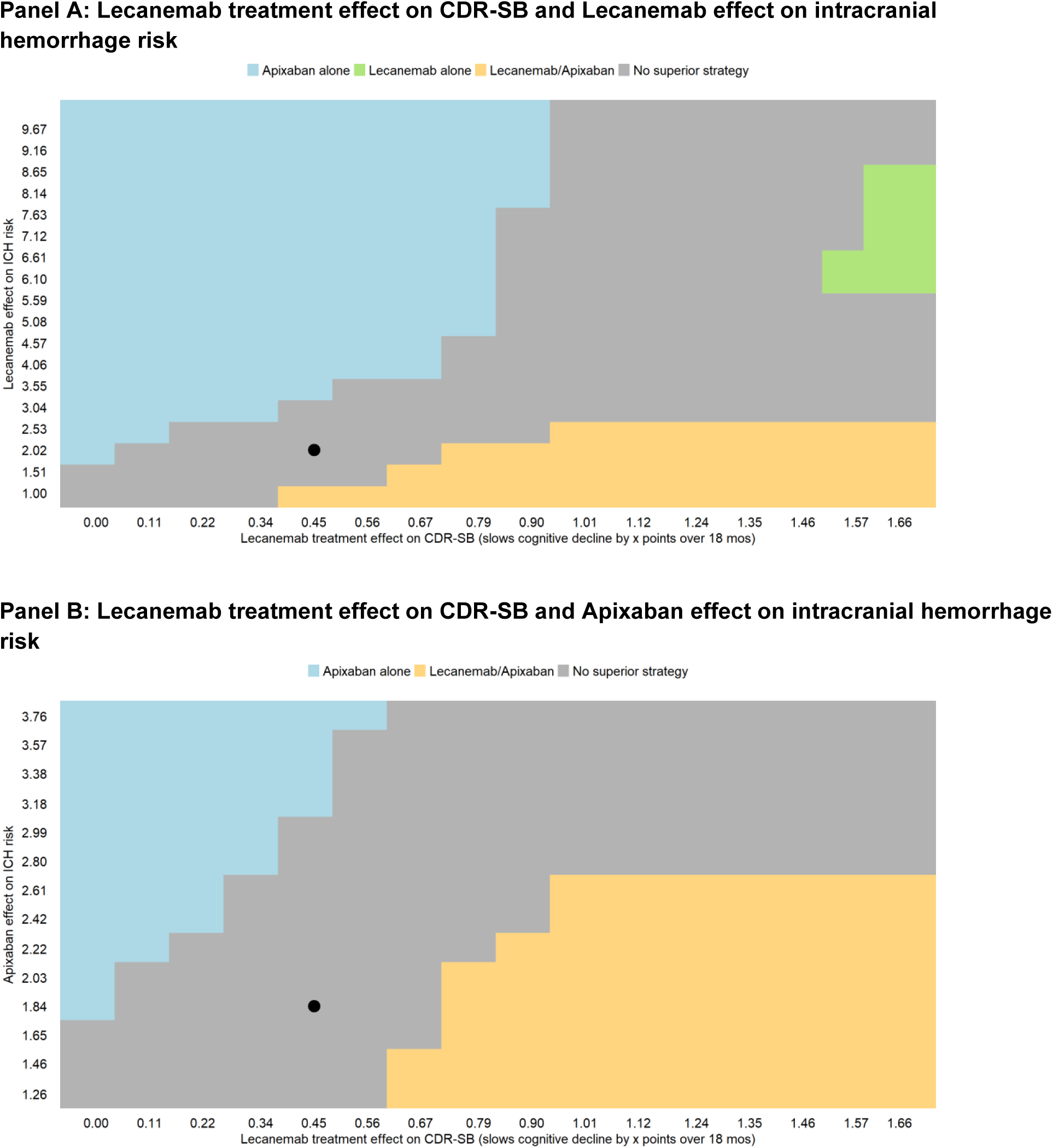
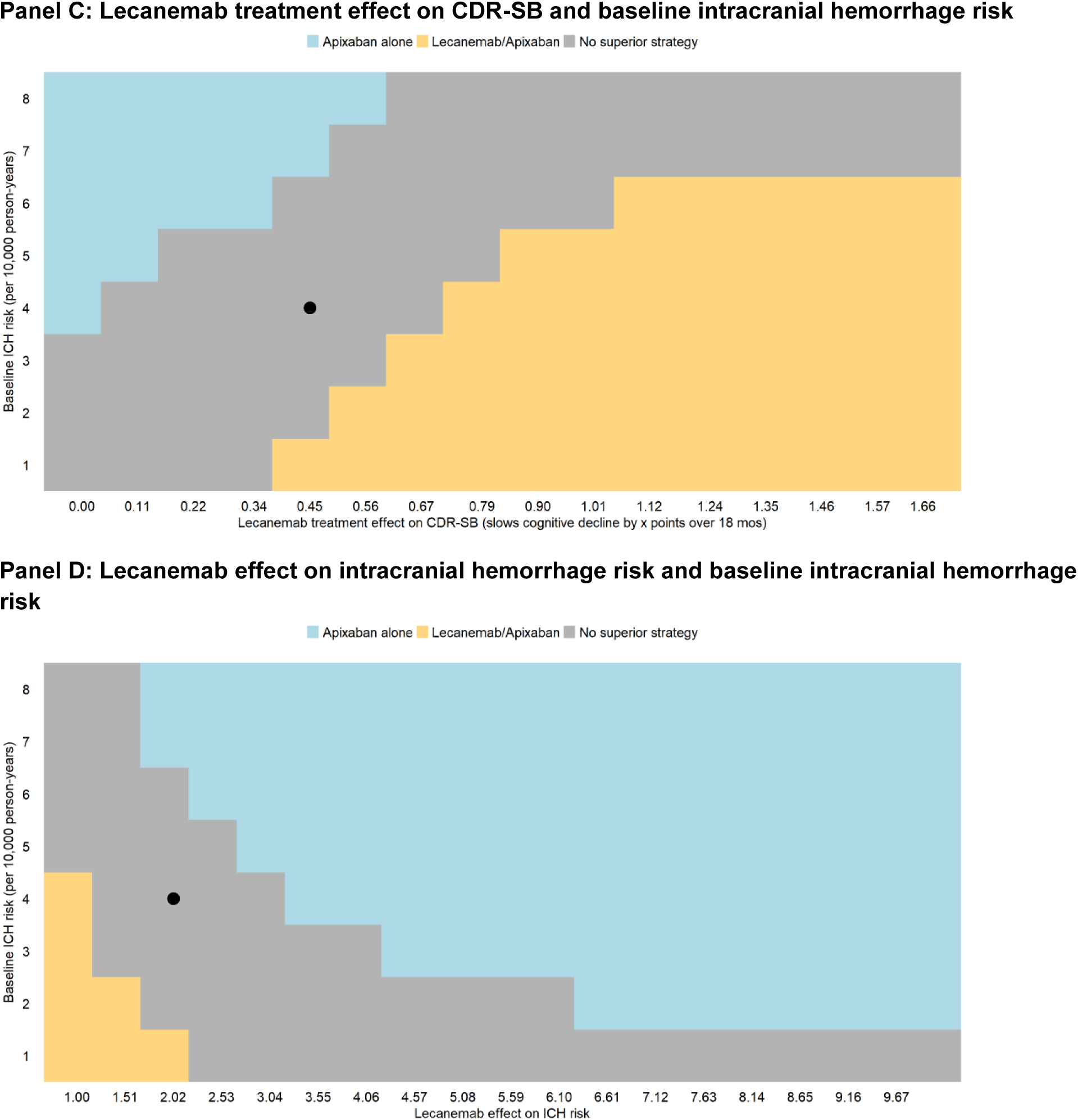
Additional model projected outcomes from multiway sensitivity analyses for people aged 65-74 years. <u>Legend</u> Multiway sensitivity analyses comparing apixaban alone (*APIX*), apixaban and lecanemab (*APIX/LEC*), and lecanemab alone (*LEC*). Panel A shows lecanemab effect on CDR-SB is varied against lecanemab effect on ICH risk. Panel B shows lecanemab effect on CDR-SB is varied against apixaban effect on ICH. Panel C shows lecanemab effect on CDR-SB is varied against baseline ICH risk. Panel D shows lecanemab effect on ICH varied against baseline ICH risk. The base case is marked by a dot. CDR-SB scores range from 0 to 18; a higher score indicates more cognitive impairment. In panels A-C, the lecanemab effect on CDR-SB as an absolute difference between untreated and treated with lecanemab; thus, higher values represent a greater effect of lecanemab. We limited precision to 0.1 quality-adjusted life months (i.e., 3 quality-adjusted life days); when the difference in QALMs of the top two strategies is less than 0.1, we display that region in gray. CDR-SB – Clinical Dementia Rating Scale Sum of Boxes; ICH – intracranial hemorrhage; QALM – quality-adjusted life months.

**Supplemental Table 1:**
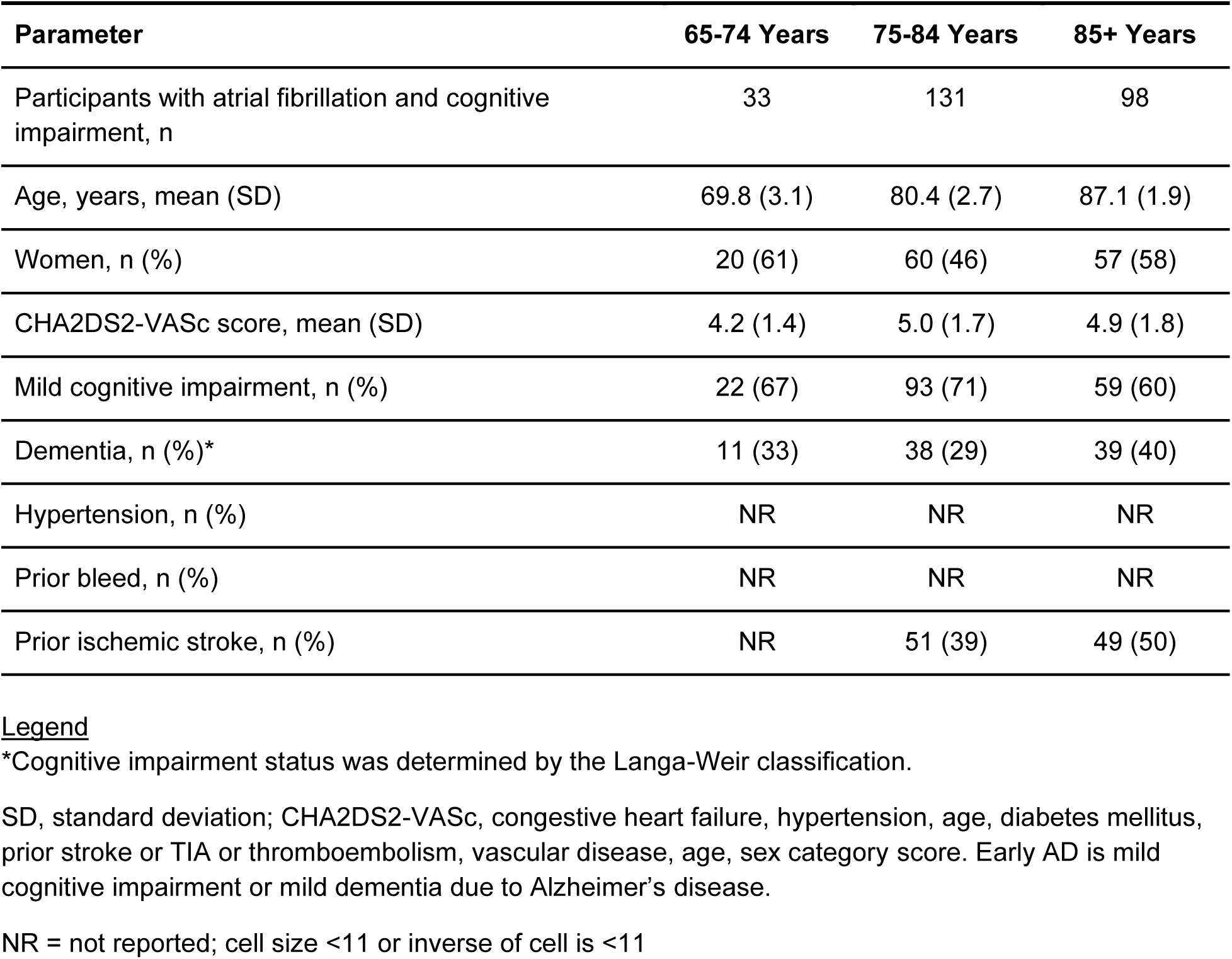
Cohort characteristics of the 2018 wave of the Health and Retirement Study who have cognitive impairment and atrial fibrillation.

| Parameter | 65-74 Years | 75-84 Years | 85+ Years |
| --- | --- | --- | --- |
| Participants with atrial fibrillation and cognitive impairment, n | 33 | 131 | 98 |
| Age, years, mean (SD) | 69.8 (3.1) | 80.4 (2.7) | 87.1 (1.9) |
| Women, n (%) | 20 (61) | 60 (46) | 57 (58) |
| CHA2DS2-VASc score, mean (SD) | 4.2 (1.4) | 5.0 (1.7) | 4.9 (1.8) |
| Mild cognitive impairment, n (%) | 22 (67) | 93 (71) | 59 (60) |
| Dementia, n (%)* | 11 (33) | 38 (29) | 39 (40) |
| Hypertension, n (%) | NR | NR | NR |
| Prior bleed, n (%) | NR | NR | NR |
| Prior ischemic stroke, n (%) | NR | 51 (39) | 49 (50) |

\*Cognitive impairment status was determined by the Langa-Weir classification.
SD, standard deviation; CHA2DS2-VASc, congestive heart failure, hypertension, age, diabetes mellitus, prior stroke or TIA or thromboembolism, vascular disease, age, sex category score. Early AD is mild cognitive impairment or mild dementia due to Alzheimer's disease.
NR = not reported; cell size <11 or inverse of cell is <11

**Supplemental Table 2:** Additional Model Input Parameters to Compare 4 Strategies for Older Adults with Early AD and Atrial Fibrillation using Apixaban (*APIX*), Apixaban and Lecanemab (*APIX/LEC*), Lecanemab (*LEC*), or Neither

| Parameter | Off<br>anticoagulant<br>probability | On<br>anticoagulant<br>probability | Reference |
| --- | --- | --- | --- |
| Health state following intracranial hemorrhage |  |  |  |
| Death (GOS 1) | 0.286 | 0.576 | (21) |
| Severe Disability (GOS 3) | 0.407 | 0.237 |  |
| Mild Disability (GOS 4) | 0.123 | 0.102 |  |
| Full recovery (GOS 5) | 0.184 | 0.085 |  |
| Health state following ischemic stroke |  |  |  |
| Death (GOS 1) | 0.117 | 0.099 | Derived from<br>(20,22,26) |
| Severe Disability (GOS 3) | 0.406 | 0.345 |  |
| Mild Disability (GOS 4) | 0.127 | 0.149 |  |
| Full recovery (GOS 5) | 0.350 | 0.408 |  |

**Supplemental Table 3:** Sensitivity analysis parameter ranges and rationale.

| Parameter | Base value | Lower range value | Upper range value | Rationale |
| --- | --- | --- | --- | --- |
| Lecanemab treatment effect on CDR-SB (CDR-SB points) | 0.45 | 0 | 1.66 | Trial reported rate is 0.45, the range is a clinically plausible effect accounting for different populations (0 equates to no effect whereas 1.66 equates to halted cognitive decline over 8 months). |
| Baseline ICH risk (per 10,000 person- year) | 4 | 8 | 1 | Represents ¼ to 2x the baseline risk; clinically plausible accounting for different populations (25) |
| Lecanemab/apixaban interaction effect on ICH risk | 2.67 | 10 | 1 | 1 represents no interaction and 10 is the top range reflecting increased uncertainty |
| Lecanemab effect on ICH risk | 2.02 | 10 | 1 | 1 represents no interaction and 10 is the top range reflecting increased uncertainty |
| Apixaban effect on ICH risk | 1.84 | 3.76 | 1.26 | 3.76 is a upper bound reported in the literature for anticoagulants (25) and 1.26 is a calculated lower bound, using a lower bound anticoagulant estimate that is further reduced to account for the safety profile of apixaban (62,63) |
| Cognitive impairment effect on mortality | 1.48 (MCI)<br>2.84 (dementia) | 1.22, 2.29 | 1.80, 3.52 | Range is based on 95% CI from Wilson 2009; note, values are smoothed between categories (33) |

Legend: MCI – mild cognitive impairment due to Alzheimer’s disease; ICH – intracranial hemorrhage; CDR-SB – Clinical Dementia Rating scale Sum of Boxes

**Supplemental Table 4:** Cognitive Impairment Quality of Life by Rating Method(36)

| **Cognitive state (CDR-SB range)** | **Caregiver proxy-rated**<br>**(base case)** | **Community-rated** | **Patient-rated** |
| --- | --- | --- | --- |
| Normal (<0.5) | 1.00 | 1.00 | 1.00 |
| MCI (0.5-4.0) | 0.80 | 0.80 | 0.86 |
| Mild dementia (4.0-9.0) | 0.74 | 0.73 | 0.85 |
| Moderate dementia (9.0-15.5) | 0.59 | 0.62 | 0.86 |

Legend: MCI – mild cognitive impairment due to Alzheimer’s disease

**Supplemental Table 5:** Scenario Analysis Comparing Strategies using Apixaban (*APIX*), Apixaban and Lecanemab (*APIX/LEC*), Lecanemab (*LEC*), or Neither using Various Source of Quality of Life Associated with Cognitive Impairment

|  | **APIX** | **APIX/LEC** | **LEC** | **Neither** |
| --- | --- | --- | --- | --- |
| **Caregiver proxy-rated quality of life (base case) (36)** |  |  |  |  |
| 65-74 years | 13.2 | 13.2 | 13.2 | 13.1 |
| 75-84 years | 12.8 | 12.6 | 12.7 | 12.6 |
| 85-90 years | 12.0 | 11.6 | 12.0 | 11.9 |
| **Community-rated quality of life (36)** |  |  |  |  |
| 65-74 years | 13.2 | 13.2 | 13.1 | 13.1 |
| 75-84 years | 12.7 | 12.5 | 12.6 | 12.6 |
| 85-90 years | 12.0 | 11.6 | 11.9 | 11.9 |
| **Patient-rated quality of life (36)** |  |  |  |  |
| 65-74 years | 14.8 | 14.7 | 14.6 | 14.6 |
| 75-84 years | 14.2 | 13.9 | 14.0 | 14.0 |
| 85-90 years | 13.5 | 13.0 | 13.3 | 13.3 |

